# Development and validation of the MMCD score to predict kidney replacement therapy in COVID-19 patients

**DOI:** 10.1101/2022.01.11.22268631

**Authors:** Flávio de Azevedo Figueiredo, Lucas Emanuel Ferreira Ramos, Rafael Tavares Silva, Magda Carvalho Pires, Daniela Ponce, Rafael Lima Rodrigues de Carvalho, Alexandre Vargas Schwarzbold, Amanda de Oliveira Maurílio, Ana Luiza Bahia Alves Scotton, Andresa Fontoura Garbini, Bárbara Lopes Farace, Bárbara Machado Garcia, Carla Thais Cândida Alves da Silva, Christiane Corrêa Rodrigues Cimini, Cíntia Alcantara de Carvalho, Cristiane dos Santos Dias, Daniel Vitorio Silveira, Euler Roberto Fernandes Manenti, Evelin Paola de Almeida Cenci, Fernando Anschau, Fernando Graça Aranha, Filipe Carrilho de Aguiar, Frederico Bartolazzi, Giovanna Grunewald Vietta, Guilherme Fagundes Nascimento, Helena Carolina Noal, Helena Duani, Heloisa Reniers Vianna, Henrique Cerqueira Guimarães, Joice Coutinho de Alvarenga, José Miguel Chatkin, Júlia Parreiras Drumond de Moraes, Juliana Machado Rugolo, Karen Brasil Ruschel, Karina Paula Medeiros Prado Martins, Luanna Silva Monteiro Menezes, Luciana Siuves Ferreira Couto, Luís César de Castro, Luiz Antônio Nasi, Máderson Alvares de Souza Cabral, Maiara Anschau Floriani, Maíra Dias Souza, Maira Viana Rego Souza e Silva, Marcelo Carneiro, Mariana Frizzo de Godoy, Maria Aparecida Camargos Bicalho, Maria Clara Pontello Barbosa Lima, Matheus Carvalho Alves Nogueira, Matheus Fernandes Lopes Martins, Milton Henriques Guimarães-Júnior, Natália da Cunha Severino Sampaio, Neimy Ramos de Oliveira, Patricia Klarmann Ziegelmann, Pedro Guido Soares Andrade, Pedro Ledic Assaf, Petrônio José de Lima Martelli, Polianna Delfino Pereira, Raphael Castro Martins, Rochele Mosmann Menezes, Saionara Cristina Francisco, Silvia Ferreira Araújo, Talita Fischer Oliveira, Thainara Conceição de Oliveira, Thaís Lorenna Souza Sales, Yuri Carlotto Ramires, Milena Soriano Marcolino

## Abstract

**Background:** Acute kidney injury (AKI) is frequently associated with COVID-19 and the need for kidney replacement therapy (KRT) is considered an indicator of disease severity. This study aimed to develop a prognostic score for predicting the need for KRT in hospitalized COVID-19 patients.

**Methods:** This study is part of the multicentre cohort, the Brazilian COVID-19 Registry. A total of 5,212 adult COVID-19 patients were included between March/2020 and September/2020. We evaluated four categories of predictor variables: (1) demographic data; (2) comorbidities and conditions at admission; (3) laboratory exams within 24 h; and (4) the need for mechanical ventilation at any time during hospitalization. Variable selection was performed using generalized additive models (GAM) and least absolute shrinkage and selection operator (LASSO) regression was used for score derivation. The accuracy was assessed using the area under the receiver operating characteristic curve (AUC-ROC). Risk groups were proposed based on predicted probabilities: non-high (up to 14.9%), high (15.0 – 49.9%), and very high risk (≥ 50.0%).

**Results:** The median age of the model-derivation cohort was 59 (IQR 47-70) years, 54.5% were men, 34.3% required ICU admission, 20.9% evolved with AKI, 9.3% required KRT, and 15.1% died during hospitalization. The validation cohort had similar age, sex, ICU admission, AKI, required KRT distribution and in-hospital mortality. Thirty-two variables were tested and four important predictors of the need for KRT during hospitalization were identified using GAM: need for mechanical ventilation, male gender, higher creatinine at admission, and diabetes. The MMCD score had excellent discrimination in derivation (AUROC = 0.929; 95% CI 0.918–0.939) and validation (AUROC = 0.927; 95% CI 0.911–0.941) cohorts an good overall performance in both cohorts (Brier score: 0.057 and 0.056, respectively). The score is implemented in a freely available online risk calculator (https://www.mmcdscore.com/).

**Conclusion:** The use of the MMCD score to predict the need for KRT may assist healthcare workers in identifying hospitalized COVID-19 patients who may require more intensive monitoring, and can be useful for resource allocation.

## BACKGROUND

Coronavirus disease 19 (COVID-19) course is mild in most cases, but it can be severe and critical, with multiple organ dysfunction, septic shock and death [1]. Kidney disease among patients with COVID-19 can manifest as acute kidney injury (AKI), hematuria, or proteinuria, and it has been associated with an increased risk of mortality [2].

The incidence of AKI among hospitalized patients with COVID-19 has shown to be variable, depending upon the severity of the disease and whether they are outpatient, in the ward or intensive care unit (ICU) environment. A recent systematic review, which included 30 studies and 18,043 patients with COVID-19, observed an overall incidence of AKI of 9.2% (95% confidence interval [CI] 4.6–13.9%), and 32.6% (95% CI 8.5–56.6%) in the ICU [3]. Another systematic review from the beginning of the pandemic included 79 studies and 49,692 patients, and observed a significant variation in the incidence of AKI and kidney replacement therapy (KRT) and the risk of death in patients who develop AKI depending on the continent. The incidence of AKI, KRT requirement and death in patients with AKI was 4.3%, 1.4% and 33.3% in Asia, 11.6%, 5.7% and 29.4% in Europe and 22.6%, 4.0% and 7.4% in North America, respectively [4]. There is a lack of studies from large cohorts in Latin America, which was severely hit by the pandemic.

Previous studies have explored the factors associated with AKI development in COVID-19 patients, including advanced age; black race; underlying medical conditions such as diabetes mellitus, cardiovascular disease, chronic kidney disease and hypertension; COVID-19 severity; use of vasopressor medications and mechanical ventilation requirement [4, 5]. However, most studies are limited to univariate analysis or have small sample sizes and there is a lack of studies analyzing independent risk factors for KRT requirement.

A risk score to predict KRT requirement during hospitalization, using clinical and laboratory data upon hospital presentation may be very useful aiming at a better allocation of health resources. However, there is a lack of evidence in this context. Fang *et al* [6] used a score created before the pandemic (UCSD-Mayo risk score) and analysed its efficiency in predicting hospital-acquired AKI in patients with COVID-19, but the performance of the score in patients in ICUs or under mechanical ventilation was not satisfactory.

Therefore, we aimed to assess the incidence of AKI and KRT requirement in COVID-19 in-hospital patients, as well as to develop and validate a score to predict the risk of the need for KRT.

## METHODOLOGY

This cohort study is part of the Brazilian COVID-19 Registry, which included consecutive patients >18 years-old, hospitalized with COVID-19 confirmed by laboratory test according to WHO criteria, admitted from March 2020 to September 2020 in 37 Brazilian hospitals, located in 17 cities, from five Brazilian states. For the present analysis, patients with chronic kidney disease stage 5 in dialysis previous to COVID-19, admitted with another diagnosis and developed COVID-19 after admission and/or those who were transferred to other hospitals (not part of the multicenter study) during hospitalization were not included. Two hospitals that did not comply with the study protocol were excluded (Figure 1).

**Figure 1.**
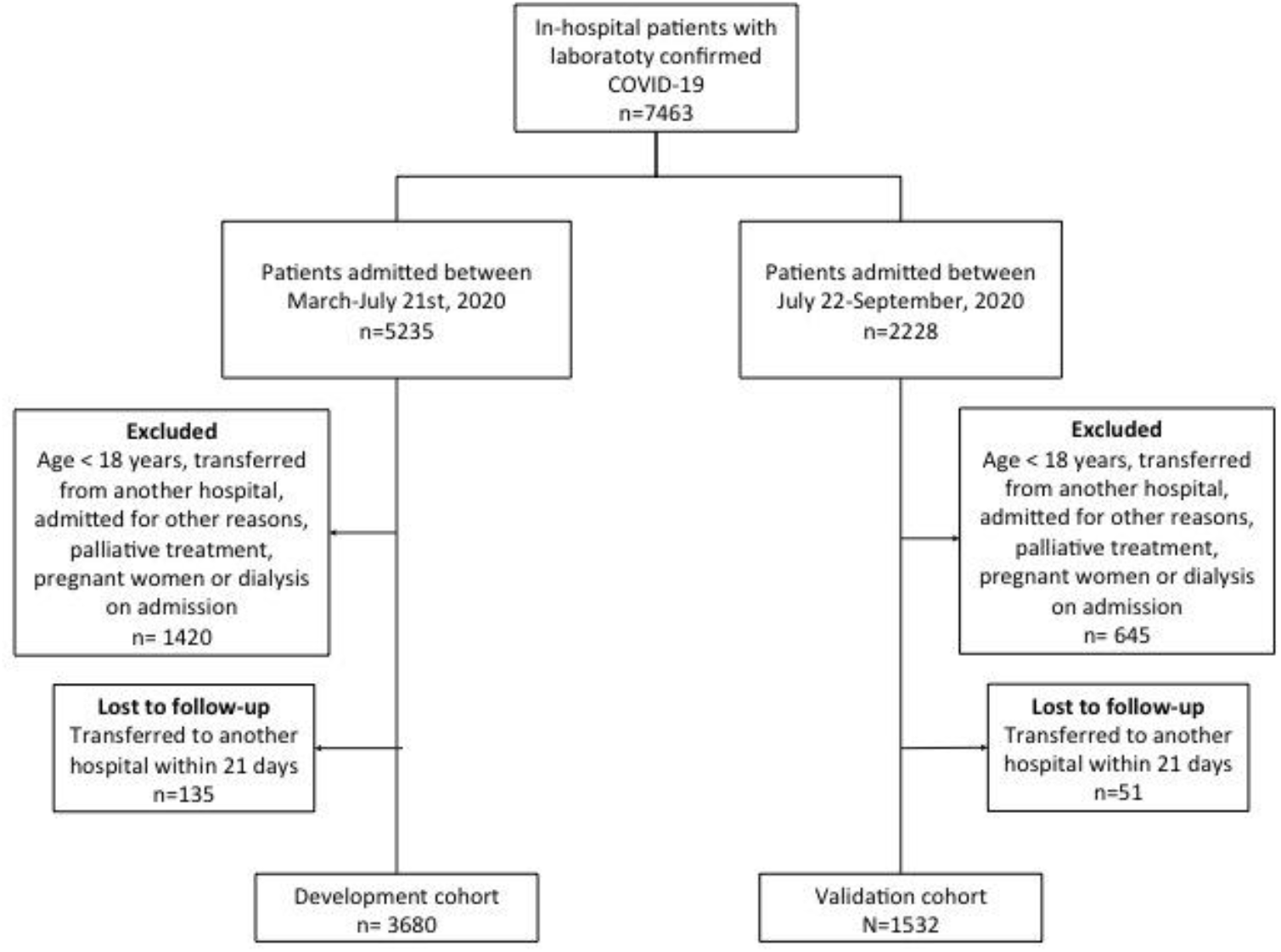
Flowchart of COVID-19 patients included in the study.

Model development, validation and reporting followed guidance from the Transparent Reporting of a Multivariable Prediction Model for Individual Prediction or Diagnosis (TRIPOD) checklist (Table S1) and Prediction model Risk Of Bias Assessment Tool (PROBAST) [7, 8].

### Data collection and outcome

Data were extracted from the medical records in participant hospitals, including patient demographic information, comorbidities, laboratory results, treatments (including KRT) and outcomes. Data were collected by using a prespecified case report form applying Research Electronic Data Capture (REDCap) tools. Variables used in the risk score were obtained at admission, with the exception of the need for mechanical ventilation, which may have occurred at any time during the hospital stay, except in those patients in which it was initiated after KRT requirement.

The primary endpoint was KRT requirement. Secondary endpoints were the incidence of AKI and mortality in patients who required KRT.

### Statistical analysis

In the descriptive analyses, categorical variables were described as absolute and relative frequency, and continuous variables by median and quartiles.

The dataset was split into development and validation, according to the date of hospital admission, using July 21, 2020 as the temporal cut (temporal validation).

All analyses were performed using R software version 4.0.2, with the mgcv, finalfit, mice, glmnet, pROC, rms, rmda, and psfmi packages. A p-value<0.05 was considered statistically significant for all analyses and 95% confidence intervals were reported.

### Missing data

Predictors were imputed if they had up to two thirds of complete values. Variables with a higher proportion of missing values than that were not included in the analysis. After analysing missing data patterns, multiple imputation with chained equations (MICE) was used to handle missing values on candidate variables, considering missing at random. Outcomes were not imputed. Predictive mean matching (PMM) method was used for imputation of continuous predictors and polytomous regression for categorical variables. The results of ten imputed datasets, each with ten iterations, were then combined, following Rubin’s rules [9].

### Development of the risk score model

Predictor selection was based on clinical reasoning and literature review before modeling, as recommended [8]. The development cohort included patients admitted before July 21, 2020.

Variable selection was performed using generalized additive models (GAM), evaluating the relationships between KRT requirement and continuous (through penalized thin plate splines) and categorical (as linear components) predictors and calculating D1-(multivariate Wald test) and D2-statistic (pools test statistics from the repeated analyses).

As our aim was to develop a score for easy application at bedside, continuous variables were categorized on cut-off points, based on evidence from an established score for sepsis [8, 10].

Subsequently, least absolute shrinkage and selection operator (LASSO) logistic regression was used to derive the score by scaling the (L1 penalized) shrunk coefficients (Table S2). Ten-fold cross-validation methods based on mean squared error criterion were used to choose the penalty parameter λ in LASSO.

Lastly, risk groups were proposed based on predicted probabilities: **non-high** (up to 14.9%), **high** (15.0 – 49.9%), and **very high** risk (≥ 50.0%).

The specific risks can be easily assessed using the developed MMCD risk score web-based calculator (https://www.mmcdscore.com), which is freely available to the public.

### Model validation

Patients who were admitted in participant hospitals from July 22, 2020 to September, 2020 were included as the external (temporal) validation cohort.

### Performance measures

To assess model calibration, predicted dialysis probabilities were plotted against the observed values. To assess model discrimination, the area under the receiver operating characteristic curve (AUROC) was calculated, with the respective confidence interval (95% CI), obtained through 2000 bootstrap samples. Positive and negative predictive values of the derived risk groups were also calculated. The Brier score was used to assess the overall performance [11].

## RESULTS

The derivation cohort included 3,680 COVID-19 patients admitted to the 35 participating hospitals, from March 1, 2020 to July 21, 2020. Those patients were from 159 cities in Brazil (Figure 2). The median age was 59 (IQR 47-70) years, 54.5% were men, 20.9% evolved with AKI, 9.3% required KRT, and 15.1% died during hospitalization. Patient demographics, underlying medical conditions, clinical characteristics, and laboratory values upon hospital presentation for the derivation and validation cohorts are displayed in Table 1.

**Table 1.** Demographic and clinical characteristics for derivation and validation cohorts of patients admitted to participant hospitals with COVID-19 (n=5,112).

| Label | Derivation cohort |  | Validation cohort |  |
| --- | --- | --- | --- | --- |
| Characteristic | N = 3,680<sup>a</sup> | Non missing cases (%) | N = 1,532<sup>a</sup> | Non missing cases (%) |
| Age (years) | 59.0 (47.0, 70.0) | 3,680 (100%) | 62.0 (48.0, 72.0) | 1,532 (100%) |
| Sex at birth |  | 3,680 (100%) |  | 1,532 (100%) |
| Men | 2,004 (54.5%) |  | 869 (56.7%) |  |
| **Comorbidities** |  |  |  |  |
| Hypertension | 1,977 (53.7%) | 3,680 (100%) | 822 (53.7%) | 1,532 (100%) |
| Coronary artery disease | 180 (4.9%) | 3,680 (100%) | 76 (5.0%) | 1,532 (100%) |
| Heart failure | 224 (6.1%) | 3,680 (100%) | 74 (4.8%) | 1,532 (100%) |
| Atrial fibrillation/flutter | 105 (2.9%) | 3,680 (100%) | 47 (3.1%) | 1,532 (100%) |
| Stroke | 106 (2.9%) | 3,680 (100%) | 53 (3.5%) | 1,532 (100%) |
| COPD | 198 (5.4%) | 3,680 (100%) | 94 (6.1%) | 1,532 (100%) |
| Diabetes mellitus | 1,009 (27.4%) | 3,680 (100%) | 453 (29.6%) | 1,532 (100%) |
| Obesity (BMI ≥ 30kg/m<sup>2</sup>) | 712 (19.3%) | 3,680 (100%) | 273 (17.8%) | 1,532 (100%) |
| Cirrhosis | 17 (0.5%) | 3,680 (100%) | 9 (0.6%) | 1,532 (100%) |
| Cancer | 170 (4.6%) | 3,680 (100%) | 75 (4.9%) | 1,532 (100%) |
| Number of Comorbidities<sup>b</sup> |  | 3,680 (100%) |  | 1,532 (100%) |
| 0 | 1,128 (30.7%) |  | 434 (28.3%) |  |
| 1 | 1,107 (30.1%) |  | 501 (32.7%) |  |
| 2 | 923 (25.1%) |  | 392 (25.6%) |  |
| 3 | 384 (10.4%) |  | 149 (9.7%) |  |
| ≥ 4 | 138 (3.8%) |  | 56 (3.7%) |  |
| **Clinical assessment at admission** |  |  |  |  |
| SF ratio | 433.3 (339.3, 452.4) | 3,582 (97%) | 433.3 (342.9, 452.4) | 1,506 (98%) |
| Heart rate (bpm) | 88.0 (78.0, 100.0) | 3,545 (96%) | 87.0 (77.0, 100.0) | 1,489 (97%) |
| Respiratory rate (irpm) | 20.0 (18.0, 24.0) | 3,043 (83%) | 20.0 (18.0, 24.0) | 1,255 (82%) |
| Glasgow coma scale | 15.0 (15.0, 15.0) | 3,460 (94%) | 15.0 (15.0, 15.0) | 1,441 (94%) |
| Systolic blood pressure |  | 3,524 (96%) |  | 1,489 (97%) |
| ≥ 90 (mm Hg) | 3,338 (94.7%) |  | 1,413 (94.9%) |  |
| < 90 (mm Hg) | 45 (1.3%) |  | 23 (1.5%) |  |
| Inotrope requirement | 141 (4.0%) |  | 53 (3.6%) |  |
| Diastolic blood pressure |  | 3,489 (95%) |  | 1,481 (97%) |
| > 60 (mm Hg) | 2,911 (83.4%) |  | 1,236 (83.5%) |  |
| ≤ 60 (mm Hg) | 437 (12.5%) |  | 192 (13.0%) |  |
| Inotrope requirement | 141 (4.0%) |  | 53 (3.6%) |  |
| Mechanical ventilation at admission | 183 (5.0%) | 3,676 (100%) | 63 (4.1%) | 1,530 (100%) |
| Mechanical ventilation after admission | 774 (21.0%) | 3,680 (100%) | 276 (18.0%) | 1,532 (100%) |
| **Laboratory parameters** |  |  |  |  |
| Hemoglobin (g/L) | 13.4 (12.2, 14.5) | 3,534 (96%) | 13.4 (12.1, 14.6) | 1,491 (97%) |
| Platelet count (109/L) | 193,500.0 (152,000.0, 252,000.0) | 3,497 (95%) | 203,000.0 (156,000.0, 263,850.0) | 1,478 (96%) |
| Neutrophils-to-lymphocytes ratio | 4.4 (2.7, 7.4) | 3,447 (94%) | 4.9 (2.9, 8.2) | 1,437 (94%) |
| Lactate value | 1.4 (1.0, 1.8) | 2,420 (66%) | 1.5 (1.1, 2.0) | 999 (65%) |
| C reactive protein (mg/L) | 71.0 (34.0, 134.6) | 3,178 (86%) | 73.5 (35.5, 134.8) | 1,311 (86%) |
| Urea (mg/dL) | 33.0 (24.0, 47.0) | 3,310 (90%) | 37.0 (27.2, 52.8) | 1,376 (90%) |
| Creatinine (mg/dL) | 0.9 (0.7, 1.2) | 3,417 (93%) | 1.0 (0.8, 1.2) | 1,434 (94%) |
| Sodium (mmol/L) | 137.0 (135.0, 140.0) | 3,215 (87%) | 137.0 (134.9, 140.0) | 1,356 (89%) |
| Bicarbonate (mEq/L) | 23.2 (21.2, 25.2) | 2,957 (80%) | 23.0 (21.0, 25.0) | 1,217 (79%) |
| pH | 7.4 (7.4, 7.5) | 2,968 (81%) | 7.4 (7.4, 7.5) | 1,217 (79%) |
| Arterial pO<sub>2</sub> | 75.0 (63.8, 94.0) | 2,927 (80%) | 74.3 (63.0, 93.7) | 1,203 (79%) |
| Arterial pCO<sub>2</sub> | 35.0 (31.9, 39.0) | 2,940 (80%) | 34.0 (30.9, 38.0) | 1,205 (79%) |
| Dialysis | 343 (9.3%) | 3,680 (100%) | 128 (8.4%) | 1,532 (100%) |
| In hospital mortality | 554 (15.1%) | 3,679 (100%) | 229 (14.9%) | 1,532 (100%) |

<sup>a</sup>Statistics presented: n (%); Median (IQR) COPD: chronic obstructive pulmonary disease, SF ratio: SpO<sub>2</sub>/FiO<sub>2</sub> ratio, BMI: body mass index. <sup>b</sup>Comorbidities included: hypertension, diabetes mellitus, obesity, coronary artery disease, heart failure, atrial fibrillation or flutter, cirrhosis, chronic obstructive pulmonary disease, cancer, and previous stroke.

**Figure 2.**
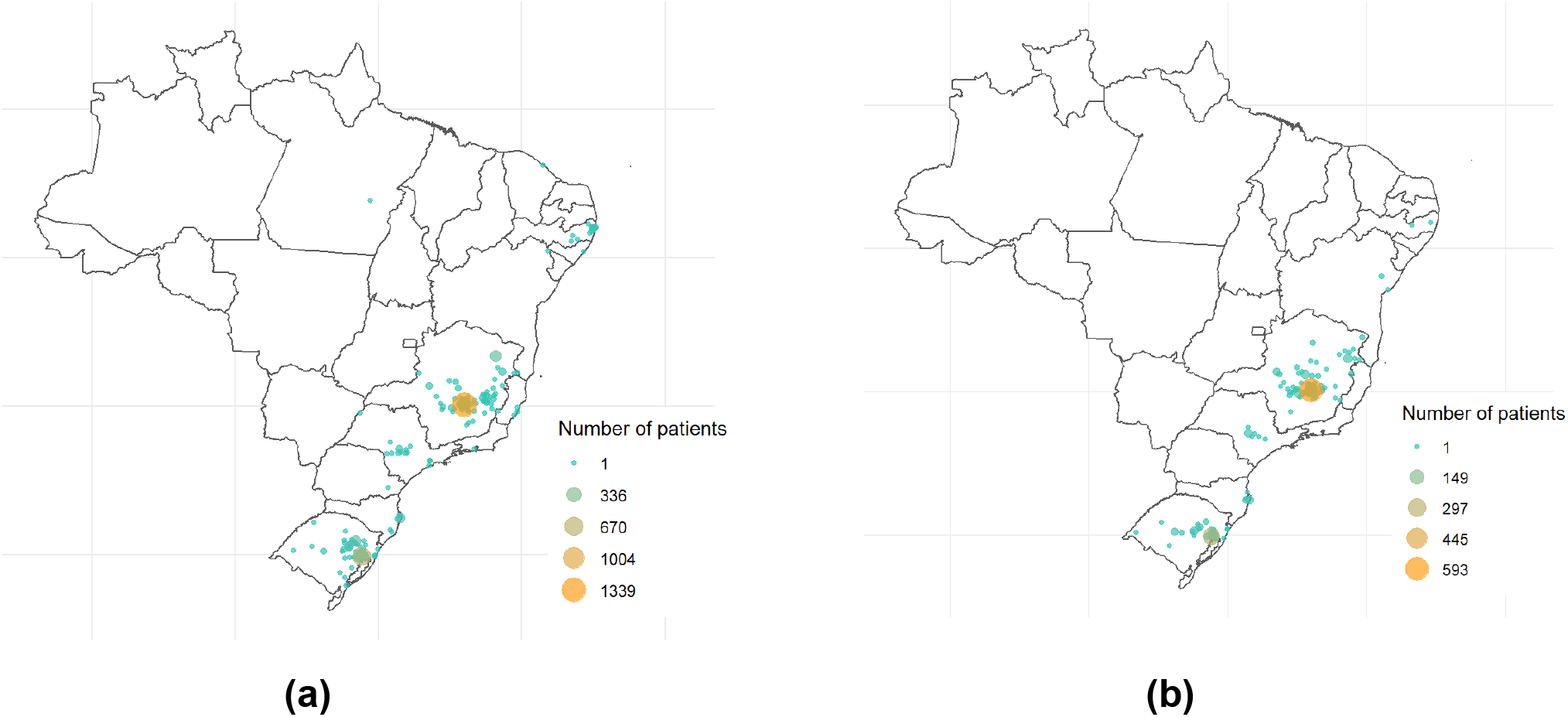
City of residence of patients from (a) the development and (b) the validation cohorts.

Among the patients in the derivation cohort, 1,261 (34.3%) required ICU admission. Of those,16.7% developed AKI and 9.1% required KRT, with a mortality rate of 64.7% and 76.7%, respectively.

### Model development

Sixty-three potential risk predictor variables collected at hospital admission were identified (Table S3). Of those, 20 were excluded for high collinearity and 11 for high number of missings variables. Consequently, 32 variables were tested.

Four important predictors of the need for KRT during hospitalization were identified using GAM: need for mechanical ventilation, male sex, higher creatinine at admission, and diabetes. Continuous selected predictors were categorized for LASSO logistic regression due to the need for a bedside use score (Table 2). The sum of the prediction scores ranges between 0 and 23, with a high score indicating higher risk of dialysis. Three risk groups were defined based on predicted probabilities of KRT requirement: non high risk (0-10 score, observed KRT rate 0.4%), high risk (11-14 score, 32.8%) and very high risk (15-23 score, 68.0%), as shown in Table 3.

**Table 2.** MMCD score for in-hospital KRT requirement in COVID-19 patients

| Variable |  | MMCD Score |
| --- | --- | --- |
| **M** | **Mechanical ventilation anytime during hospital stay<sup>a</sup>** |  |
|  | No | 0 |
|  | Yes | 11 |
| **M** | **Sex** |  |
|  | Women | 0 |
|  | Men | 1 |
| **C** | **Creatinine (mg/dL) upon hospital presentation** |  |
|  | < 1.2 | 0 |
|  | 1.2 - 2.0 | 1 |
|  | 2.0 - 3.5 | 2 |
|  | 3.5 - 5.0 | 4 |
|  | ≥ 5.0 | 10 |
| **D** | **Diabetes mellitus** |  |
|  | No | 0 |
|  | Yes | 1 |

<sup>a</sup> Except in those cases the dialysis preceded mechanical ventilation.

**Table 3.** Predicted probability of dialysis and dialysis rates for MMCD score risk groups

| Risk Groups |  | Derivation cohort |  | Validation cohort |  |
| --- | --- | --- | --- | --- | --- |
| Risk Group | Predicted probability | No. of patients | No. of dialysis cases | No. of patients | No. of dialysis cases |
| Non high risk (0-10) | 0 - 14.9% | 2,719 | 10 (0.4%) | 1,192 | 8 (0.7%) |
| High (11-14) | 15 - 49.9% | 911 | 299 (32.8%) | 310 | 108 (34.8%) |
| Very high (15-23) | ≥ 50% | 50 | 34 (68%) | 30 | 12 (40%) |
| Overall | - | 3,680 | 343 (9.3%) | 1,532 | 128 (8.4%) |

Discrimination and model overall performance in derivation and validation cohorts for GAM, LASSO and MMCD score are shown in Table 4. Within the derivation cohort, the MMCD risk score showed excellent discrimination (AUROC = 0.929; 95% CI 0.918–0.939) a good overall performance (Brier score: 0.057) (figure 3a).

**Table 4.** Discrimination and model overall performance in derivation and validation cohorts

| Models | Derivation cohort |  | Validation cohort |  |
| --- | --- | --- | --- | --- |
| Model | AUROC (95% CI) | Brier score | AUROC (95% CI) | Brier score |
| GAM | 0.938 (0.926-0.947) | 0.053 | 0.917 (0.893-0.937) | 0.057 |
| LASSO | 0.929 (0.918-0.938) | 0.057 | 0.929 (0.914-0.943) | 0.055 |
| MMCD score | 0.929 (0.918-0.939) | 0.057 | 0.927 (0.911-0.941) | 0.056 |

**Figure 3.**
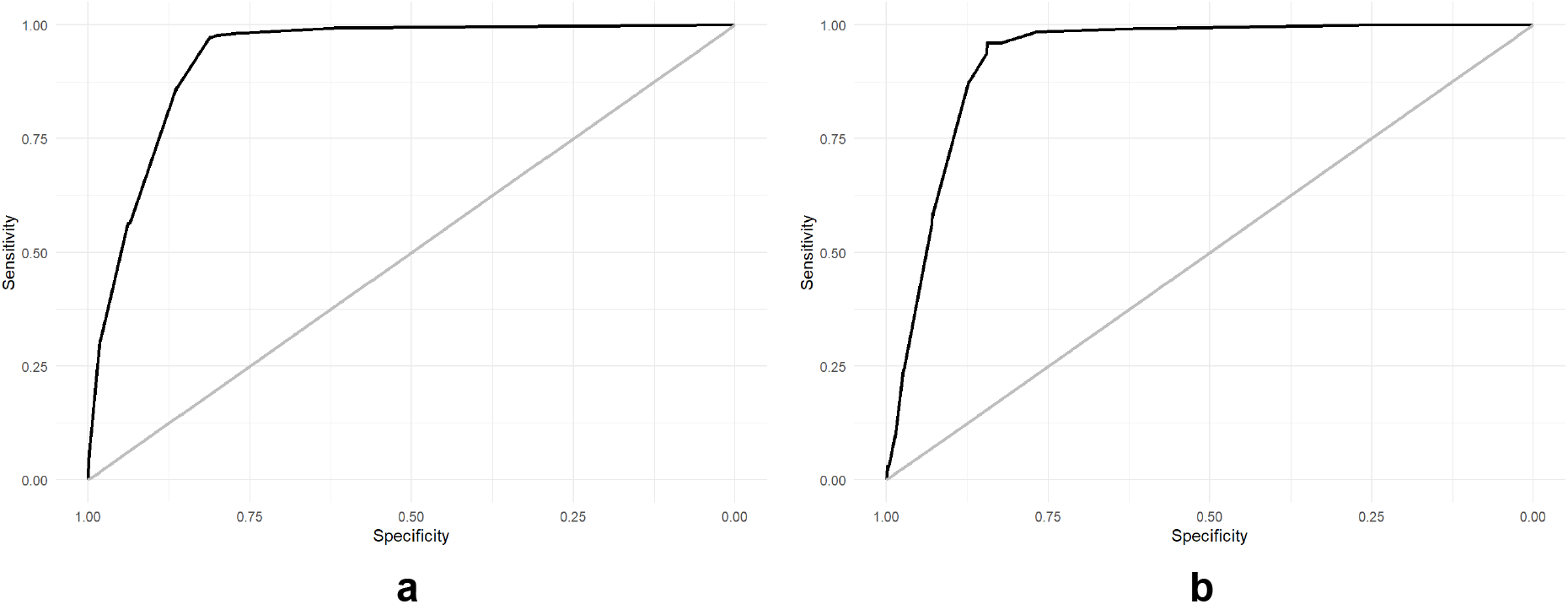
Derivation (a) and Validation (b) cohort ROC curve.

### Model validation

A total of 1,532 patients admitted between July 22, 2020 and September 31, 2020 were included in the validation cohort. The median age was 62 (IQR 48-72) years; 56.7% were male, 19.8% evolved with AKI, 8.4% required KRT, and 14.9% died during hospitalization.

In the validation cohort, 515 (33.6%) required ICU admission. Of those, 14.6% developed AKI and 8.1% required KRT, with a mortality rate of 65.5% and 82.3%, respectively.

The MMCD Score had an AUROC of 0.927 (95% CI 0.911-0.941), calibration and performance were good (slope=0.849, Brier score=0.056 intercept=-0.186), under the validation cohort (Figure 3b, Figure S1 and Figure S2).

## DISCUSSION

This study included more than 5,000 patients hospitalized with a diagnosis of COVID-19 in the different hospitals included in the Brazilian COVID-19 Registry, a robust cohort of patients with a comprehensive dataset. One in every five patients evolved with AKI and 9.3% required KRT. Among the analyzed predictors, four variables were related to progression to AKI and KRT requirement, including: the need for mechanical ventilation, sex, creatinine upon hospital presentation and diabetes mellitus.

Renal involvement in COVID-19 infection is complex and probably occurs due to several factors. Direct renal injury by the SARS-CoV-2 to the renal endothelium, tubular epithelium and podocytes has been described, infecting the cell through angiotensin converting enzyme 2 receptors, which are abundantly present in the renal tissue [12]. In addition to this mechanism, there are numerous factors worsening the kidney injury, such as: cytokine storm with the release of several interleukins and cytokines, mainly interleukin-6 (IL-6) [3]; cardiorenal syndrome, caused by right ventricular dysfunction secondary to pulmonary infection; hypercoagulable state with a coronary lesion, left ventricular dysfunction and consequent low renal output, worsened by hypovolemia; and release of nephrotoxic substances such as creatine phosphokinase secondary to rhabdomyolysis [2].

The need for mechanical ventilation at any time during hospitalization was an important predictor of progression to AKI and the need for KRT, being the variable with the highest points in the risk score. Scoring mechanical ventilation only changed patients’ category to “high risk” for evolving to AKI and KRT requirement. Another study from the USA (United States of America) with 5,449 patients, the need for mechanical ventilation was a significant risk factor for AKI and KRT requirement [5]. This finding confirms findings from studies carried out in other countries, such as a study in the USA from the beginning of the pandemic, in which mechanical ventilation was the strongest predictor for AKI (OR 10.7 [95% CI 6.81–16.70]) [5]. In another study from the United Kingdom, which included ICU patients, mechanical ventilation was also the strongest predictor, associated with a four-fold risk of progression of AKI in COVID-19 patients [13]. Study in patients hospitalized with COVID-19 in the city of Wuhan, China, with a significant difference in the need for mechanical ventilation between patients with and without AKI (54.4% vs. 13.7%) [14]. Finally, a study developed in Brazil with patients hospitalized in a single-center with COVID-19 that described the need for mechanical ventilation among patients without AKI, with AKI and with AKI associated with KRT needed were 17.0%, 69,3% and 100% [15]. There is a close relationship between alveolar and tubular damage (lung-kidney axis) in acute respiratory distress syndrome (ARDS), often progressing to different degrees of AKI [16]. The relationship between mechanical ventilation (MV) and AKI has been widely recognized before the COVID-19 pandemic. Animal models suggest a causal relationship between MV and AKI through a reduction in renal blood flow, apparently by an impairment in intrarenal microcirculation due to hypoxemia and hypercapnia, as well as a drop in cardiac output due to changes in intrathoracic pressure with MV [17]. Additionally, the extrinsic positive end expiratory pressure (PEEP) seems to be associated with a redistribution of intra-renal blood flow, and “biotrauma” may be another cause. This is a complex and not fully understood mechanism, in which inflammatory mediators are released by ventilated lungs into the systemic circulation [18].

Therefore, AKI in patients who require mechanical ventilation seems to be multifactorial, and it is difficult to define the specific role that each mechanism plays in the pathogenesis of AKI. They are usually observed simultaneously in critically ill patients, which limits the possibility to develop preventive strategies [19].

In studies published by Chan L et al (n=3,993) [20] and Fisher M et al (n=3,345) [21] with hospitalized patients with COVID-19 in the USA, male sex was considered an independent predictor of progression to AKI and KRT requirement. In our study, the male sex was a risk predictor variable for the evolution of AKI and the need for KRT, being included in the risk score. Male sex has been previously observed to be associated with other adverse outcomes in COVID-19 patients. In a recent meta-analysis with over three million COVID-19 cases, the authors observed no difference in the proportion of men and women who developed COVID-19, but men had almost three times the odds of requiring ICU admission (OR = 2.84; 95% CI = 2.06, 3.92) and higher odds of death (OR = 1.39; 95% CI = 1.31, 1.47) compared to women [22].

Creatinine levels upon hospital presentation may be evidence of previous chronic kidney disease or an early manifestation of AKI caused by COVID-19 infection. Chronic kidney disease is a global health problem and a silent disease [23]. Serum creatinine levels were categorized according to the Sequential Organ Failure Assessment Score (SOFA) [10] to comply with TRIPOD guidelines, which advises not to use a data-driven method, to avoid model overfitting [8]. Our finding is consistent with a recent systematic review and meta-analysis with 22 studies (n=17,391), which observed an increased incidence of AKI in COVID-19 patients hospitalized in the USA who had abnormal baseline serum creatinine levels due to pre-existing chronic kidney disease [24]. Hansrivijit P et al [25] in their meta-analysis described abnormal basal serum creatinine levels as predictors of progression to AKI.

The association between diabetes mellitus and renal dysfunction is well known, in the form of diabetic nephropathy, as a result not only of intrarenal atherosclerosis and arteriosclerosis, but also non inflammatory glomerular damage [26, 27]. Among the predictor variables analyzed in this study, diabetes proved to be a predictor of risk of progression to AKI and KRT requirement in patients hospitalized with COVID-19. A meta-analysis published in 2020 with 26 studies (n=5,497) evaluated the incidence of AKI in patients with COVID-19 and showed that diabetes was a predisposing factor for progression to AKI [25].

Meta-analyses showed that the association of patients diagnosed with COVID-19 who developed AKI had higher mortality, which was enhanced by the need for KRT [4, 28]. In Brazil, a country severely hit by the pandemic, there is lack of evidence on the association among AKI, need for KRT, mortality and COVID-19. The scarce existing studies are based in small databases. A study published with 200 ICU-patients showed a high incidence of AKI (about 50%) and 17% of patients requiring KRT, with significantly higher mortality in patients with AKI and needing KRT, in contrast to patients without AKI and KRT requirement [15]. In our study, the incidence of AKI and need for KRT in ICU-patients were lower (about 16% and 9%, respectively), although with higher in-hospital death in this group, similarly to finds in this article.

The MMCD model retrieved an AUROC of 0.96, which was classified as a excellent discrimination. An American study (n=2,256) developed prediction models of KRT using machine learning techniques, comparing L1-penalized logistic regression (logistic L1), elastic-net logistic regression (logistic EN) and gradient boosted trees (GBT). Logistic L1 had the best accuracy in the validation cohort. However, the discrimination results were inferior than the one observed in the present analysis (0.847 [95% CI, 0.772-0.936]) and the study has several limitations: many risk predictor variables, hindering the applicability of the score and high incidence of missing variables [29].

### Strengths and limitations

Our study used a large patients database to develop a risk score to predict the need for KRT in patients admitted with COVID-19. A major strength of the MMCD score is its simplicity; the use of objective parameters, which may reduce the variability; and easy availability, even in under-resourced settings. Then, the MMCD score may help clinicians to make a prompt and reasonable decision to optimize the management of COVID-19 patients with AKI and potentially reduce mortality. Additionally, our article strictly followed the TRIPOD recommendations [8].

This study has limitations. Indication and timing of initiation of the KRT may differ according to institutional protocols, and we did not collect information on patients who did not perform dialysis due to limited resources. Still, this has not affected the accuracy of the score. Additionally, as any other score, MMCD may not be directly generalized to populations from other countries. Furthermore, it was not possible to use the KDIGO (Kidney Disease: Improving Global Outcomes) classification for AKI due to the lack of data on previous serum creatinine of patients admitted to participating hospitals. Instead, we used the SOFA score, which has been widely used for ICU patients for years, and it is currently recommended to assess organ dysfunction in patients suspected of sepsis [30]. In patients with abnormal serum creatinine levels, it was not possible to define the causal factor (previous chronic kidney disease vs. COVID-19) due to the lack of data prior to the patient’s hospitalization.

### Possible applications

Using predictors available at baseline and within the first hours of the admission, we could objectively predict the probability of KRT of a COVID-19 patient with AKI. With an accurate prediction, it may help to organize resource allocation to patients who are at the highest risk of KRT requirement [29], in addition to selecting patients who may benefit from renal protection strategies, close assessment and follow-up by a nephrologist [31].

## CONCLUSIONS

In conclusion, we developed and validated a clinical prediction score named MMCD, to predict the need for KRT in COVID-19 patients. This score used a few predictors available at baseline and mechanical ventilation anytime during hospital admission, and retrieved a good accuracy. This could be an inexpensive tool to predict the need for KRT objectively and accurately. Additionally, it may be used to inform clinical decisions and the assignment to the appropriate level of care and treatment for COVID-19 patients with AKI.

## Data Availability

All data produced in the present study are available upon reasonable request to the authors

## List of abbreviations

AKI: acute kidney injury
ARDS: acute respiratory distress syndrome
AUROC: area under the receiver operating characteristic
BMI: body mass index
BPM1: beats per minute
BPM2: breaths per minute
CI: confidence interval
COPD: chronic obstructive pulmonary disease
COVID-19: Coronavirus disease 19
GAM: generalized additive models
GBT: gradient boosted trees
ICU: intensive care unit
IL-6: interleukin-6
IQR: interquartile range
KDIGO: Kidney Disease Improving Global Outcomes
KRT: kidney replacement therapy
LASSO: least absolute shrinkage and selection operator
MICE: multiple imputation with chained equations
MMCD: mechanical ventilation, male, creatinine, diabetes
MV: mechanical ventilation
pCO2: partial pressure of carbon dioxide
PEEP: positive end-expiratory pressure
PMM: Predictive mean matching
PO2: partial pressure of oxygen
PROBAST: Prediction model Risk Of Bias Assessment Tool
REDCap: Research Electronic Data Capture
SBP: systolic blood pressure
SF ratio: SpO2/FiO2 ratio
SOFA: Sepsis-related Organ Failure Assessment
TRIPOD: Transparent Reporting of a Multivariable Prediction Model for Individual Prediction or Diagnosis
USA: United States of America

## DECLARATIONS

### Ethics approval and consent to participate

This study was approved by Brazilian National Committee for Ethics in Research - CONEP (CAAE - Certificate of presentation of Ethical Appreciation: 30350820.5.1001.0008).

### Consent for publication

Informed consent was waived due to the pandemic situation and the study design, based on data collection from medical records only.

### Availability of data and materials

Any additional data pertaining to this manuscript are available from the corresponding author upon reasonable request.

### Competing interests

The authors declare that they have no competing interests. The sponsors had no role in study design; data collection, management, analysis, and interpretation; writing the manuscript; and deciding to submit it for publication.

### Funding

This study was supported in part by Minas Gerais State Agency for Research and Development (*Fundação de Amparo à Pesquisa do Estado de Minas Gerais - FAPEMIG*) [grant number APQ-00208-20] and National Institute of Science and Technology for Health Technology Assessment (*Instituto de Avaliação de Tecnologias em Saúde – IATS*)/ National Council for Scientific and Technological Development (*Conselho Nacional de Desenvolvimento Científico e Tecnológico - CNPq*) [grant number 465518/2014-1].

### Authors’ contributions

Conception or design of the work: MSM and MCP. Data collection: AVS, AOM, ALBAS, AFG, BLF, BMG, CTCAS, CCRC, CAC, DVS, EFRM, EPAC, FA, FGA, FCA, FB, GGV, GFN, HCN, HD, HRV, HCG, JCA, JMC, JPDM, JMR, KBR, KPMPM, LSMM, LSFC, LCC, LAN, MASC, MAF, MDS, MVRSS, MC, MFG, MACB, MCPBL, MCAN, MFLM, MHGJ, NCSS, NRO, PKZ, PGSA, PLA, PJLM, PDP, RLRC, RCM, RMM, SCF, SFA, TFO, TCO, TLSS, YCR and MSM. Data analysis and interpretation: MSM, MCP, LEFR, RTS and FAF. Drafting the article: FAF, MSM, MCP, CSD and DP. Critical revision of the article: all authors. Final approval of the version to be published: all authors.

## Acknowledgements

We would like to thank the hospitals, which are part of this collaboration: Hospital das Clínicas da Faculdade de Medicina de Botucatu; Hospital Universitário de Santa Maria; Hospital São João de Deus; Hospital Regional Antônio Dias; Hospital Nossa Senhora da Conceição; Hospital Cristo Redentor; Hospital Risoleta Tolentino Neves; Hospital Júlia Kubitschek; Hospital Santo Antônio; Hospital Santa Rosália; Hospital João XXIII; Hospital UNIMED BH; Hospital Mãe de Deus; Hospital Universitário Canoas; Hospital SOS Cárdio; Hospital das Clínicas da Universidade Federal de Pernambuco; Hospital das Clínicas da UFMG; Hospital Universitário Ciências Médicas; Hospital São Lucas da PUCRS; Hospital Luxemburgo; Hospital Metropolitano Odilon Behrens; Hospital Moinhos de Vento; Hospital Bruno Born; Hospital Santa Cruz; Hospitais da Rede Mater Dei; Hospital Márcio Cunha; Hospital Eduardo de Menezes; Hospital Tacchini; Hospital Semper and Hospital Metropolitano Doutor Célio de Castro. We also thank all the clinical staff at those hospitals, who cared for the patients, and all students who helped with the data collection.

## Notes

### Competing Interest Statement

The authors have declared no competing interest.

