## Supplementary material for "Development and validation of the MMCD score to predict kidney replacement therapy in COVID-19 patients"

Article

**Supplementary material**

Table S1. TRIPOD checklist for transparent reporting on a multivariable prognostic model ………………………………………………………………………………….….... p.2

Table S2. L1 penalized shrunk coefficients for the MMCD score……….………….... p.4

Table S3. Assessment of potential predictors for the model development …………. p.5

Figure S1. Calibration slope for the MMCD score …...………………………………... p.7

Figure S2. Combined decision curve for the MMCD score …………...………...….... p.7

References…………………………………………………………………….………….... p.8

**Table S1.** TRIPOD checklist for transparent reporting on a multivariable prognostic model.

| **Section/Topic** | **Item** |  | **Checklist Item** | **Page** |
| --- | --- | --- | --- | --- |
| **Title and abstract** | | | | |
| Title | 1 | D;V | Identify the study as developing and/or validating a multivariable prediction model, the target population, and the outcome to be predicted. | 1 |
| Abstract | 2 | D;V | Provide a summary of objectives, study design, setting, participants, sample size, predictors, outcome, statistical analysis, results, and conclusions. | 9 |
| **Introduction** | | | | |
| Background and objectives | 3a | D;V | Explain the medical context (including whether diagnostic or prognostic) and rationale for developing or validating the multivariable prediction model, including references to existing models. | 10-11 |
|  | 3b | D;V | Specify the objectives, including whether the study describes the development or validation of the model or both. | 12 |
| **Methods** | | | | |
| Source of data | 4a | D;V | Describe the study design or source of data (e.g., randomized trial, cohort, or registry data), separately for the development and validation data sets, if applicable. | 12 |
|  | 4b | D;V | Specify the key study dates, including start of accrual; end of accrual; and, if applicable, end of follow-up. | 12 |
| Participants | 5a | D;V | Specify key elements of the study setting (e.g., primary care, secondary care, general population) including number and location of centres. | 12-13 |
|  | 5b | D;V | Describe eligibility criteria for participants. | 12 |
|  | 5c | D;V | Give details of treatments received, if relevant. | N.A. |
| Outcome | 6a | D;V | Clearly define the outcome that is predicted by the prediction model, including how and when assessed. | 13 |
|  | 6b | D;V | Report any actions to blind assessment of the outcome to be predicted. | N.A. |
| Predictors | 7a | D;V | Clearly define all predictors used in developing or validating the multivariable prediction model, including how and when they were measured. | 19, Tab.2 |
|  | 7b | D;V | Report any actions to blind assessment of predictors for the outcome and other predictors. | N.A. |
| Sample size | 8 | D;V | Explain how the study size was arrived at. | N.A. |
| Missing data | 9 | D;V | Describe how missing data were handled (e.g., complete-case analysis, single imputation, multiple imputation) with details of any imputation method. | 14 |
| Statistical analysis methods | 10a | D | Describe how predictors were handled in the analyses. | 14,15, 16 |
|  | 10b | D | Specify type of model, all model-building procedures (including any predictor selection), and method for internal validation. | 15 |
|  | 10c | V | For validation, describe how the predictions were calculated. | 17-18 |
|  | 10d | D;V | Specify all measures used to assess model performance and, if relevant, to compare multiple models. | 15 |
|  | 10e | V | Describe any model updating (e.g., recalibration) arising from the validation, if done. | N.A. |
| Risk groups | 11 | D;V | Provide details on how risk groups were created, if done. | 15, 16, 17 |
| Development vs. validation | 12 | V | For validation, identify any differences from the development data in setting, eligibility criteria, outcome, and predictors. | 15, 16, 17 |
| **Results** | | | | |
| Participants | 13a | D;V | Describe the flow of participants through the study, including the number of participants with and without the outcome and, if applicable, a summary of the follow-up time. A diagram may be helpful. | 12, 13 Fig. 1 |
|  | 13b | D;V | Describe the characteristics of the participants (basic demographics, clinical features, available predictors), including the number of participants with missing data for predictors and outcome. | 15, 16 Tab. 1 |
|  | 13c | V | For validation, show a comparison with the development data of the distribution of important variables (demographics, predictors and outcome). | 18, 19 Tab. 1 |
| Model development | 14a | D | Specify the number of participants and outcome events in each analysis. | 15,16 |
|  | 14b | D | If done, report the unadjusted association between each candidate predictor and outcome. | N.A. |
| Model specification | 15a | D | Present the full prediction model to allow predictions for individuals (i.e., all regression coefficients, and model intercept or baseline survival at a given time point). | Tab. S2 |
|  | 15b | D | Explain how to the use the prediction model. | 17, Tab.2 |
| Model performance | 16 | D;V | Report performance measures (with CIs) for the prediction model. | 18, Tab.4 |
| Model-updating | 17 | V | If done, report the results from any model updating (i.e., model specification, model performance). | N.A. |
| **Discussion** | | | | |
| Limitations | 18 | D;V | Discuss any limitations of the study (such as nonrepresentative sample, few events per predictor, missing data). | 24 |
| Interpretation | 19a | V | For validation, discuss the results with reference to performance in the development data, and any other validation data. | 19 Fig.3 |
|  | 19b | D;V | Give an overall interpretation of the results, considering objectives, limitations, results from similar studies, and other relevant evidence. | 20-23 |
| Implications | 20 | D;V | Discuss the potential clinical use of the model and implications for future research. | 24 |
| **Other information** | | | | |
| Supplementary information | 21 | D;V | Provide information about the availability of supplementary resources, such as study protocol, Web calculator, and data sets. | 15 |
| Funding | 22 | D;V | Give the source of funding and the role of the funders for the present study. | 27 |

**Table S2.** L1 penalized shrunk coefficients for the MMCD score

|  | **Variable** | **Coefficient** |
| --- | --- | --- |
| **M** | **Intercept**  **Mechanical ventilation anytime during hospital stay^a^** | -4,841 |
|  | No | - |
|  | Yes | 3,682 |
| **M** | **Sex** |  |
|  | Women | - |
|  | Men | 0,213 |
| **C** | **Creatinine (mg/dL) upon hospital presentation** |  |
|  | < 1.2 | - |
|  | 1.2 - 2.0 | 0,306 |
|  | 2.0 - 3.5 | 0,755 |
|  | 3.5 - 5.0 | 1,444 |
|  | ≥ 5.0 | 3,179 |
| **D** | **Diabetes mellitus** |  |
|  | No | - |
|  | Yes | 0,263 |

**^a^** Except in those cases the dialysis preceded mechanical ventilation.

**Table S3:** Assessment of potential predictors for the model development

| **Variables** | **Scientific evidence** | **Model development (derivation cohort)** |
| --- | --- | --- |
| **Demographic** |  |  |
| Age in years (continuous) | [1] [2] [3] [4] [5] [6] [7] [8] [9] [10] | Included as candidate predictor |
| Sex at birth | [2] [3] [6] [7] [8] [9] [10] | Included as candidate predictor |
| **Conditions highly and moderately associated with increased risk of complications (NHS guidance)** | - | - |
| Cardiovascular system | - | - |
| - Hypertension | [2] [5] [6] [7] [8] [10] | Included as candidate predictor |
| - Coronary artery disease | [2] [3] [5] [6] [7] [8] [10] | Included as candidate predictor |
| - Heart failure | [2] [5] [6] [7] [8] [10] | Included as candidate predictor |
| - Atrial fibrillation/flutter | [3] [5] [8] [10] | Included as candidate predictor |
| - Ischemic stroke | [10] | Included as candidate predictor |
| Diabetes mellitus^[1]^ | [2] [3] [5] [6] [7] [8] [10] | Included as candidate predictor |
| Obesity (BMI>30kg/m2) | [3] [5] [7] [8] [10] | Included as candidate predictor |
| Cirrhosis | [5] [8] [10] | Included as candidate predictor |
| Chronic kidney disease | [2] [3] [8] [9] [10] | Included as candidate predictor |
| HIV infection | [3] [8] [10] | Included as candidate predictor |
| Malignant neoplasm | [2] [3] [5] [7] [8] | Included as candidate predictor |
| Previous transplantation | [10] | Included as candidate predictor |
| Report of hospital surgical procedure in the last 90 days |  | Included as candidate predictor |
| **Lifestyle** |  | Included as candidate predictor |
| Illicit drugs use |  | Included as candidate predictor |
| Alcohol abuse |  | Included as candidate predictor |
| Current smoking | [7] [8] [9] [10] | Included as candidate predictor |
| Previous smoking | [7] [10] | Included as candidate predictor |
| **Clinical findings** | - | - |
| Symptoms time | [7] | Included as candidate predictor |
| Sensory impairment | [10] | High collinearity with Invasive Mechanical Ventilation, not included |
| Glasgow come scale | [10] | High collinearity with Invasive Mechanical Ventilation, not included |
| Mental status | [10] | - |
| - Alert |  | High collinearity with SBP, not included |
| - Confused |  | High collinearity with SBP, not included |
| - Disoriented |  | High collinearity with SBP, not included |
| - Sleepy |  | High collinearity with SBP, not included |
| - Torporous |  | High collinearity with SBP, not included |
| - Coma |  | High collinearity with SBP, not included |
| Systolic blood pressure (mmHg) | [3] [4] [9] [10] | Included as candidate predictor |
| Diastolic blood pressure (mmHg) | [3] [4] [9] [10] | High collinearity with SBP, not included |
| Use of vasoactive amines | [3] [4] [7] [8] [9] [10] | High collinearity with SBP, not included |
| Heart rate (bpm) | [3] [9] [10] | Included as candidate predictor |
| Respiratory rate (bpm) | [3] [9] [10] | Included as candidate predictor |
| O2 saturation (%) | [3] [8] [9] [10] | Included as candidate predictor |
| Invasive Mechanical Ventilation at admission | [3] [6] [7] [8] [9] [10] | Included as candidate predictor |
| Invasive Mechanical Ventilation on third day of hospitalization | [3] [6] [7] [8] [9] | High collinearity with Invasive Mechanical Ventilation, not included |
| Invasive Mechanical Ventilation on fifith day of hospitalization | [3] [6] [7] [8] [9] | High collinearity with Invasive Mechanical Ventilation, not included |
| Invasive Mechanical Ventilation at any time of hospitalization | [3] [6] [7] [8] [9] | High collinearity with Invasive Mechanical Ventilation, not included |
| **Laboratory findings** | - | - |
| Hemoglobin (g/dL) | [3] [7] [8] [9] [10] | High collinearity with NLR, not included |
| Leukocytes (cells/mm3) | [2] [5] [6] [7] [8] [9] | High collinearity with NLR, not included |
| Neutrophils (cels/mm3) | [2] [5] [6] [8] [9] | High collinearity with NLR, not included |
| Lymphocytes (cels/mm3) | [2] [3] [5] [6] [7] [8] [9] | High collinearity with NLR, not included |
| NLR | [2] [3] [5] [6] | Included as candidate predictor |
| Platelets (cels/mm3) | [2] [3] [6] [7] [8] [9] [10] | High collinearity with NLR, not included |
| Albumin (g/dL) | [3] [5] [6] [9] [10] | Too many missing values, not included |
| Creatinine (mg/dL) | [3] [4] [6] [7] [8] [9] | Included as candidate predictor |
| Creatine phosphokinase (CPK - U/L) | [2] [7] [9] | Too many missing values, not included |
| D-dimer (ng/ml) | [6] [7] [9] [10] | Too many missing values, not included |
| Ferritin (ng/mL) | [7] [8] [9] [10] | Too many missing values, not included |
| Lactate dehydrogenase (LDH) (U/L) | [7] [9] | Too many missing values, not included |
| Protein C reactive (mg/L) | [2] [3] [6] [8] [9] [10] | Included as candidate predictor |
| Procalcitonin (ng/mL) | [6] [7] [9] [10] | Too many missing values, not included |
| PTTa (seconds)/control | [9] | Too many missing values, not included |
| RNI | [9] | Too many missing values, not included |
| Sodium (mmoL) | [5] [7] [8] [9] | Included as candidate predictor |
| TGO/AST (U/L) | [3] [6] [7] [8] [9] | Too many missing values, not included |
| TGP/ALT (U/L) | [3] [5] [6] [7] [8] [9] | Too many missing values, not included |
| Troponin | [9] [10] | Too many missing values, not included |
| Urea (mg/dL) | [3] [7] [8] [9] [10] | Included as candidate predictor |
| pH | [7] [8] [9] | Included as candidate predictor |
| arterial pCO2 | [7] [9] | Included as candidate predictor |
| arterial pO2 | [3] [7] [8] [9] [10] | High collinearity with pH, not included |
| HCO3- | [3] [7] [8] [9] | High collinearity with pCO2, not included |

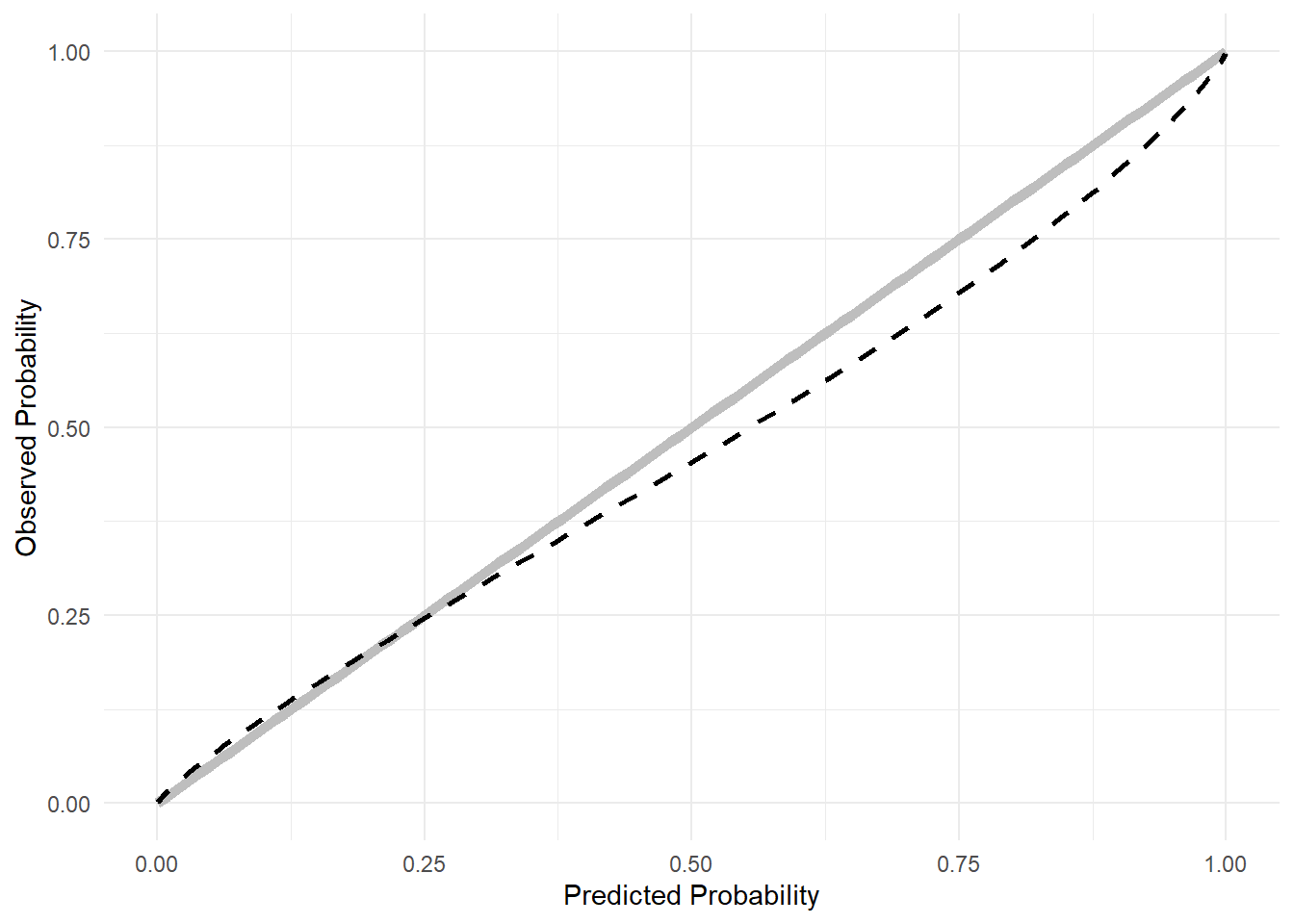

**Figure S1:** Calibration slope for the MMCD score

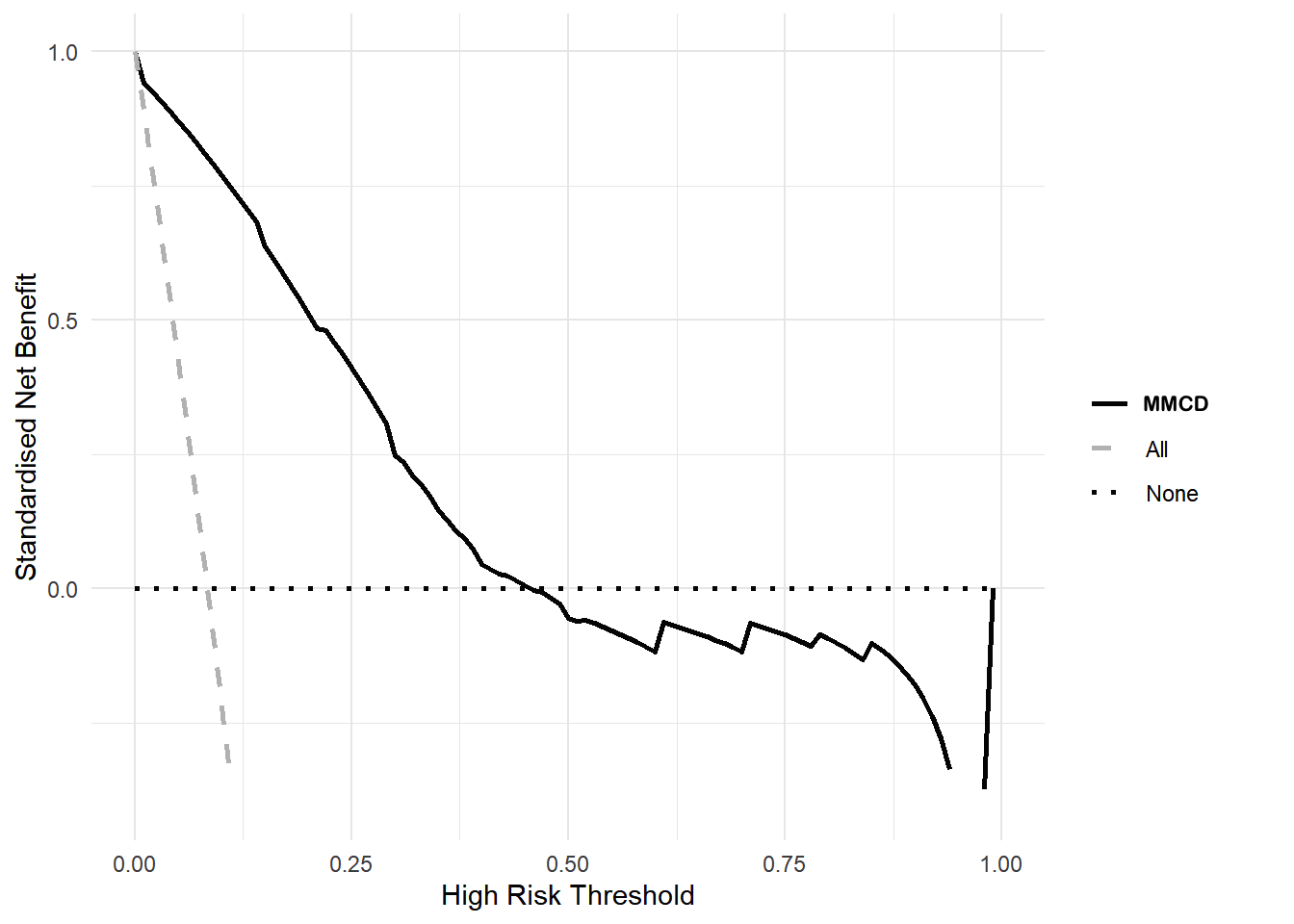

**Figure S2.** Combined decision curve for the MMCD score
